# Natural aphrodisiacs consumption by male workers in the former Katanga province, DR Congo

**DOI:** 10.1101/2024.02.07.24302074

**Authors:** Paul Musa Obadia, Gaylord Kalenda Mulaji, Taty Muta Musambo, Joseph Pyana Kitenge, Trésor Carsi Kuhangana, Tony Kayembe-Kitenge, Célestin Banza Lubaba Nkulu, Benoit Nemery, Paul Enzlin

## Abstract

**Background:** In a previous cross-sectional study to determine the prevalence and determinants of erectile dysfunction (ED) among artisanal mineworkers, we found a significant association between ED and consumption of locally produced plant-derived aphrodisiacs.

**Aim:** We aimed to investigate the type and mode of consumption of aphrodisiacs, their possible health effects, and reasons for use among male workers in the Katanga province.

**Methods:** We conducted a mixed method study, first doing a survey (March 2021) among a convenience sample of 127 artisanal miners and 122 collective taxi-drivers. Participants responded to the International Index of Erectile Function (IIEF) questionnaire and had measurements of serum urea, creatinine, hepatic enzymes, total cholesterol, high-density lipoprotein (HDL), low-density lipoprotein (LDL), triglycerides and sexual hormones. In a second qualitative phase, 16 participants, i.e., eight miners and eight drivers were interviewed.

**Results:** Self-reported regular (at least once a week) consumption of aphrodisiacs was significantly more prevalent among taxi-drivers (75%) than among miners (47%). Mild-to-moderate and moderate ED were reported more frequently by aphrodisiacs consumers [20/152 (13%)] than non-consumers [6/97 (6%)]. Five types of plant-derived aphrodisiacs (*Zingiber officinale, Mitragyna stipulosa, Ocimum* sp*, Piper nigrum, Securidaca longepedunculata*) were consumed, via the oral or anal routes, sometimes together with alcohol. We found no evidence of nephrotoxicity, hepatotoxicity or disturbances in serum levels of sex hormones. Themes that emerged from interviews related to cultural perceptions about masculinity, with miners using aphrodisiacs to support failing erectile function, and taxi-drivers taking aphrodisiacs for preventing ED.

**Conclusion:** We found a high prevalence of plant-derived aphrodisiacs use among working men in Katanga. While no serious adverse effects were identified, more ethno-botanical studies with isolation and rigorous identification of active constituents are needed to provide the poor population with correct information and to protect them against possible unwanted toxic effects.

## Introduction

For centuries and throughout the world, men have felt the need to take products to support erectile function [1]. Many plants or animals are known or believed to contain compounds capable of enhancing erectile function. Such natural products to improve erectile function are called “aphrodisiacs” in reference to Aphrodite, the Greek goddess of love and beauty. Aphrodisiacs can be categorized into three types according to their mode of action: i.e., libido increasing, sexual pleasure increasing, and potency increasing [2,3].

In spite of the availability of pharmacologically approved drugs to treat erectile dysfunction (ED), i.e., the phosphodiesterase type 5 (PDE5) inhibitors sildenafil, tadalafil, vardenafil, and avanafil [4], aphrodisiac plants remain widely used in Africa, where herbal-based treatments play an important role in the treatment and management of communicable and non-communicable diseases [5]. In most African communities, medicinal plants are regarded as accessible, affordable, and acceptable [5]. This is in line with a recent report from the World Health Organization (WHO) stating that more than 80% of the population in Africa still continue to rely on traditional medicines for their primary health care [6]. Similarly, a recent study of perceptions and use of traditional African medicine in Lubumbashi [Democratic Republic of Congo (DRC)], showed that 80% of participants reported using traditional medicines, which was defined as herbal-based treatments [7]. Studies conducted in Kinshasa [8] and in Lubumbashi [9] reported a large number of plants that are used to treat ED and prescribed mostly by less educated traditional healers.

In a previous cross-sectional study to determine the prevalence of ED among artisanal mineworkers, we found a significant association between aphrodisiacs consumption and erectile dysfunction [10]. Here, we report on a mixed methods follow-up study investigating the use of aphrodisiacs by male workers in Katanga. In a first phase, consisting of a cross-sectional survey among mineworkers and taxi drivers, we used questionnaires to assess erectile function and the prevalence of taking aphrodisiacs, as well as their types, origin and modes of use; we also measured sex hormones and markers of liver or kidney damage in serum. This was followed by a second qualitative phase in which we used interviews to explore the reasons why these men used aphrodisiacs.

## 2. Methods

We conducted a mixed methods study [11,12] using an explanatory sequential design in which a quantitative phase (survey) was followed by a qualitative phase (interviews).

### Phase I. Quantitative survey

In a cross-sectional survey (23/03/2021 - 28/05/2021), we included, by convenience sampling at their workplace, 127 artisanal miners working in three copper-cobalt mines (Kamilombe, Tshabula and UCK) in Kolwezi, and 122 drivers of collective taxis (minibuses, called “matatu” in East Africa) based in Lubumbashi but sometimes working on the Lubumbashi– Kolwezi line. Miners included in the present study were different from our previous study on the prevalence of ED [10] but they belong to the same groups as those included in a study on hypoxemia during underground work [13].

As described in our previous survey [10], we used a questionnaire to obtain sociodemographic and other relevant characteristics including age (sometimes only an approximate date of birth), marital status, number of children, age of the last child, extra conjugal partner, current and past residence, occupation, lifestyle (tobacco smoking, alcohol consumption, drug use), current or chronic illnesses (diabetes, hypertension), use of medication, including PDE5 inhibitors, and current consumption of local aphrodisiacs (at least once a week). To assess male sexual function, we used a Swahili version [10] of the International Index of Erectile Function (IIEF) questionnaire, which provides scores for five domains (erectile function, orgasmic function, sexual desire, satisfaction with intercourse, and general satisfaction) and a total score of male sexual function [14]. The IIEF has been shown to have good psychometric quality, and the internal consistency in the current sample was excellent (Cronbach’s alpha = 0.95) [15]. ED was defined as a score for erectile function (IIEF-EF) <25, with the following degrees of severity: mild ED (score 19-24), mild-to-moderate (score 13-18), moderate (score 7-12) and severe (score <7).

The paper questionnaires were generally self-administered, but for participants with low literacy face-to-face assistance was sometimes needed. Reportedly consumed aphrodisiac plants were later identified from their vernacular name by botanists from the *Institut National d’Etudes et Recherches Agronomiques* (INERA) KIPOPO in Lubumbashi.

While the participant was seated, blood pressure was measured using an Omron M3 HEM-7131-IntelliSense Blood Pressure Monitor. Height and weight were measured with a rod and a SECA scale, respectively. The body mass index (BMI) also was calculated.

At the end of the interview, in participants consenting with blood sampling, blood was drawn from a brachial vein between 7 AM and 11 AM (after thorough cleaning and disinfection of the skin with alcohol) into a 5 mL BD Vacutainer with spray-coated SSTTM II Advance (Becton Dickinson REF 369032) to obtain serum. Urea, creatinine, sGOT, sGPT, total cholesterol, HDL, LDL, triglyceride were measured using the CYANSmart Semi-Automatic Biochemistry Analyser CYPRESS DIAGNOSTICS (Hulshout, Belgium). Serum concentrations of testosterone, Sex Hormone Binding Globulin (SHBG) and Luteinizing Hormone (LH) were measured, as previously [13], by competitive immunoassay using ECLIA technology on a COBAS e801 instrument (Roche Diagnostics). Free testosterone was calculated with the Vermeulen formula [16].

### Statistical analysis

Statistical analyses were conducted using SPSS (IBM SPSS Statistics for Windows, Version 26.0.Armonk, NY) and Prism 10.2 (GraphPad Software, San Diego, CA 9210, USA) for graphical illustration. Statistical significance was defined as a two-tailed p-value ≤0.05. Distribution of quantitative data was tested with the Shapiro-Wilk test. Normally distributed data are presented as means with their standard deviation (SD). Non-parametric data are reported as medians with their interquartile range (IQR). Differences between groups were assessed for statistical significance by Student’s t-test, Mann Whitney U-test or Kruskall-Wallis test, as appropriate, and for proportions by the Fisher’s Exact Test. Logistic regression will be used to check the association between aphrodisiacs consumption and blood tests, and socio demographic parameters.

### Phase II: Qualitative phase: One-to-one Interviews

After completion of the survey, we interviewed some participants to explore the reasons why they took aphrodisiacs. Based on purposive sampling, we invited eight taxi-drivers and eight miners who had declared using currently local aphrodisiac products. After obtaining informed consent, the same person (PMO) conducted one-to-one interviews in Swahili at a time and place convenient to the participants. Interviews lasted for about 10 minutes and were audio-recorded, with field notes taken simultaneously. Sufficient prompts were utilized to obtain in-depth information, for additional clarification and to avoid being deviated from the topic. Debriefing was done at the end of each interview. The interview was transcribed (by PMO). The final report was rechecked and given back to the participants to confirm its accuracy. Transcripts of audio-taped interviews were translated into French, and analysed by a single person (PMO) using thematic analysis [17]. In the results section, quotes are attributed to individual respondents, designated as M for miners and T for taxi-drivers + respondent number. The IDs mentioned after each quote are only known by the interviewer (PMO).

### Ethical aspects

The study protocol was approved by the Committee for medical Ethics of the Université de Lubumbashi (UNILU). Approval number: UNILU/CEM /144 /2018. All respondents included in the study signed a written consent form.

## 3. Results

### 3.1. General characteristics

Table 1 presents sociodemographic and other relevant characteristics of the 249 participants, i.e., 127 miners and 122 taxi-drivers. The two groups had a similar mean age. No participant reported a history of diabetes or hypertension. More taxi-drivers than miners reported drinking alcohol, smoking tobacco and using aphrodisiacs. Self-reported aphrodisiacs consumption at least once a week was 1.6-fold more prevalent among taxi-drivers (75%) compared to miners (47%). Obesity was infrequent (3%), but more prevalent among taxi drivers. High blood pressure (systolic pressure ≥140 mmHg or diastolic pressure ≥90 mmHg) was present in 16% taxi-drivers and in 3% of miners.

**Table 1:**
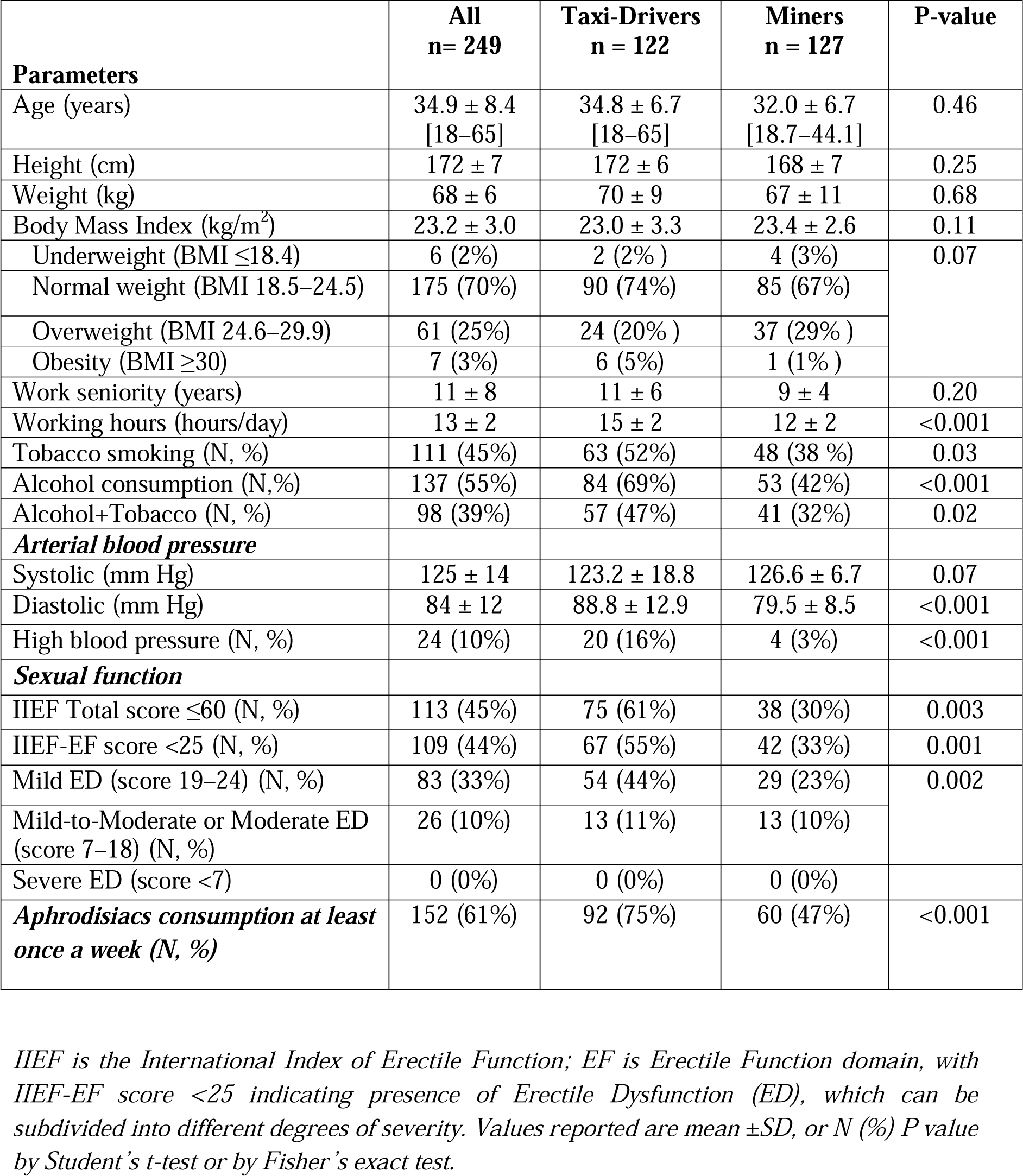
Sociodemographic and other characteristics of participants.

| Parameters | All<br>n= 249 | Taxi-Drivers<br>n = 122 | Miners<br>n = 127 | P-value |
| --- | --- | --- | --- | --- |
| Age (years) | 34.9 ± 8.4<br>[18–65] | 34.8 ± 6.7<br>[18–65] | 32.0 ± 6.7<br>[18.7–44.1] | 0.46 |
| Height (cm) | 172 ± 7 | 172 ± 6 | 168 ± 7 | 0.25 |
| Weight (kg) | 68 ± 6 | 70 ± 9 | 67 ± 11 | 0.68 |
| Body Mass Index (kg/m<sup>2</sup>) | 23.2 ± 3.0 | 23.0 ± 3.3 | 23.4 ± 2.6 | 0.11 |
| Underweight (BMI ≤18.4) | 6 (2%) | 2 (2%) | 4 (3%) | 0.07 |
| Normal weight (BMI 18.5–24.5) | 175 (70%) | 90 (74%) | 85 (67%) |  |
| Overweight (BMI 24.6–29.9) | 61 (25%) | 24 (20%) | 37 (29%) |  |
| Obesity (BMI ≥30) | 7 (3%) | 6 (5%) | 1 (1%) |  |
| Work seniority (years) | 11 ± 8 | 11 ± 6 | 9 ± 4 | 0.20 |
| Working hours (hours/day) | 13 ± 2 | 15 ± 2 | 12 ± 2 | <0.001 |
| Tobacco smoking (N, %) | 111 (45%) | 63 (52%) | 48 (38%) | 0.03 |
| Alcohol consumption (N,%) | 137 (55%) | 84 (69%) | 53 (42%) | <0.001 |
| Alcohol+Tobacco (N, %) | 98 (39%) | 57 (47%) | 41 (32%) | 0.02 |
| **Arterial blood pressure** |  |  |  |  |
| Systolic (mm Hg) | 125 ± 14 | 123.2 ± 18.8 | 126.6 ± 6.7 | 0.07 |
| Diastolic (mm Hg) | 84 ± 12 | 88.8 ± 12.9 | 79.5 ± 8.5 | <0.001 |
| High blood pressure (N, %) | 24 (10%) | 20 (16%) | 4 (3%) | <0.001 |
| **Sexual function** |  |  |  |  |
| IIEF Total score ≤60 (N, %) | 113 (45%) | 75 (61%) | 38 (30%) | 0.003 |
| IIEF-EF score <25 (N, %) | 109 (44%) | 67 (55%) | 42 (33%) | 0.001 |
| Mild ED (score 19–24) (N, %) | 83 (33%) | 54 (44%) | 29 (23%) | 0.002 |
| Mild-to-Moderate or Moderate ED (score 7–18) (N, %) | 26 (10%) | 13 (11%) | 13 (10%) |  |
| Severe ED (score <7) | 0 (0%) | 0 (0%) | 0 (0%) |  |
| **Aphrodisiacs consumption at least once a week (N, %)** | 152 (61%) | 92 (75%) | 60 (47%) | <0.001 |

*IIEF is the International Index of Erectile Function; EF is Erectile Function domain, with* *IIEF-EF score <25 indicating presence of Erectile Dysfunction (ED), which can be* *subdivided into different degrees of severity. Values reported are mean ±SD, or N (%) P value* *by Student's t-test or by Fisher's exact test.*

With regards to sexual function as assessed by the IIEF, questionnaire-defined ED (IIEF-EF score <25) was more prevalent among taxi drivers (55%) than among artisanal miners (34%). This was mainly due to a higher prevalence of mild ED (IIEF score 19–24) among taxi drivers, because the proportions of men with mild-to-moderate ED were similar in both groups. Only one man (a taxi-driver) had a score below 12 suggestive of moderate ED, and none of the participants had severe ED (score <7). The proportion of regular consumers of aphrodisiacs was higher among taxi-drivers (75%) than miners (47%).

Table 2 presents sociodemographic and other relevant characteristics of men according to the use of aphrodisiacs. The 152 regular consumers (36.6 ± 8.6y) were on average 4 years older than the 97 non-consumers (32.0 ± 7.4y); obesity was more prevalent among aphrodisiacs consumers. Reported alcohol drinking, tobacco smoking, and cannabis use were more prevalent among aphrodisiacs consumers than non-consumers. High blood pressure was measured in 15% of consumers and in 2% of non-consumers. The prevalence rates of any ED (IIEF score <25) did not differ between regular users (41%) and non-users of aphrodisiacs (48%). However, mild-to-moderate or moderate ED was more prevalent among regular users (13%) than non-users (6%).

**Table 2:** Characteristics of participants according to regular use of aphrodisiacs.

| Parameters | Regular consumers<br>n = 152 | Non-consumers<br>n = 97 | P-value |
| --- | --- | --- | --- |
| Age (years) | 36.6 ± 8.6<br>[19 – 65] | 32.4 ± 7.4<br>[17.8 – 55] | <0.001 |
| Height (cm) | 172 ± 7 | 171 ± 7 | 0.1 |
| Weight (kg) | 69 ± 11 | 66 ± 11 | 0.06 |
| Body Mass Index (kg/m<sup>2</sup>) | 23.1 ± 3.2 | 23.2 ± 2.7 | 0.87 |
| Underweight (BMI ≤18.4) | 4 (3%) | 2 (2%) | 0.28 |
| Normal weight (BMI 18.5–24.5) | 110 (72%) | 65 (67%) |  |
| Overweight (BMI 24.6–29.9) | 32 (21%) | 29 (30%) |  |
| Obesity (BMI ≥30) | 6 (4%) | 1 (1%) |  |
| Work seniority (years) | 12 ± 8 | 9 ± 6 | 0.01 |
| Working hours (hours/day) | 13 ± 3 | 13 ± 2 | 0.23 |
| Tobacco smoking (N, %) | 89 (59%) | 22 (23%) | <0.001 |
| Alcohol consumption (N, %) | 104 (68%) | 33 (34%) | <0.001 |
| Alcohol+Tobacco (N, %) | 78 (51%) | 20 (21%) | <0.001 |
| Cannabis | 71 (47%) | 23 (24%) | <0.001 |
| **Arterial blood pressure** |  |  |  |
| Systolic (mm Hg) | 125 ± 16 | 124 ± 12 | 0.49 |
| Diastolic (mm Hg) | 87 ± 13 | 80 ± 9 | <0.001 |
| High blood pressure (N, %) | 22 (15%) | 2 (2%) | 0.001 |
| **Sexual function** |  |  |  |
| IIEF total score ≤60 (N, %) | 70 (46%) | 43 (44%) | 0.79 |
| IIEF-EF <25 (N, %) | 62 (41%) | 47 (48%) | 0.24 |
| Mild ED (score 19–24) | 42 (28%) | 41 (42%) | 0.022 |
| Mild-to-Moderate or Moderate (score 7–18) (N, %) | 20 (13%) | 6 (6%) |  |
| Severe ED (score <7) | 0 (0%) | 0 (0%) |  |

*IIEF is the International Index of Erectile Function; EF is Erectile Function domain, with* *IIEF-EF score <25 indicating presence of Erectile Dysfunction (ED), which can be* *subdivided into different degrees of severity. Values reported are mean ± SD or N (%); P* *value by Student's t-test or by Fisher's exact test.*

### 3.2. Aphrodisiacs consumption

As shown in Figure 1, traditional aphrodisiac products are easily available in the area.

**Figure 1:**
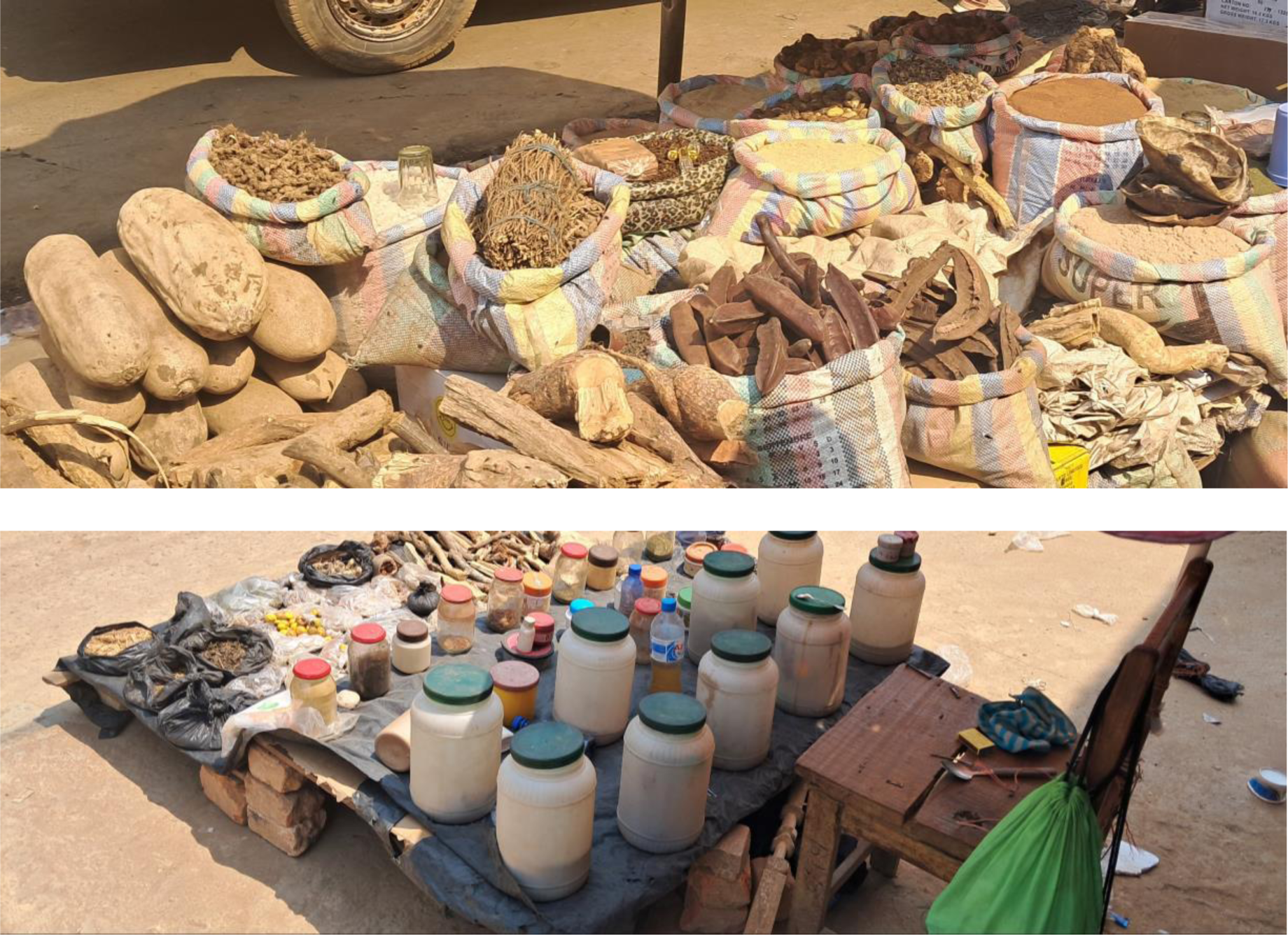
Traditional medicine products (including aphrodisiacs) as sold publicly in Lubumbashi down town.

**Figure 2.**
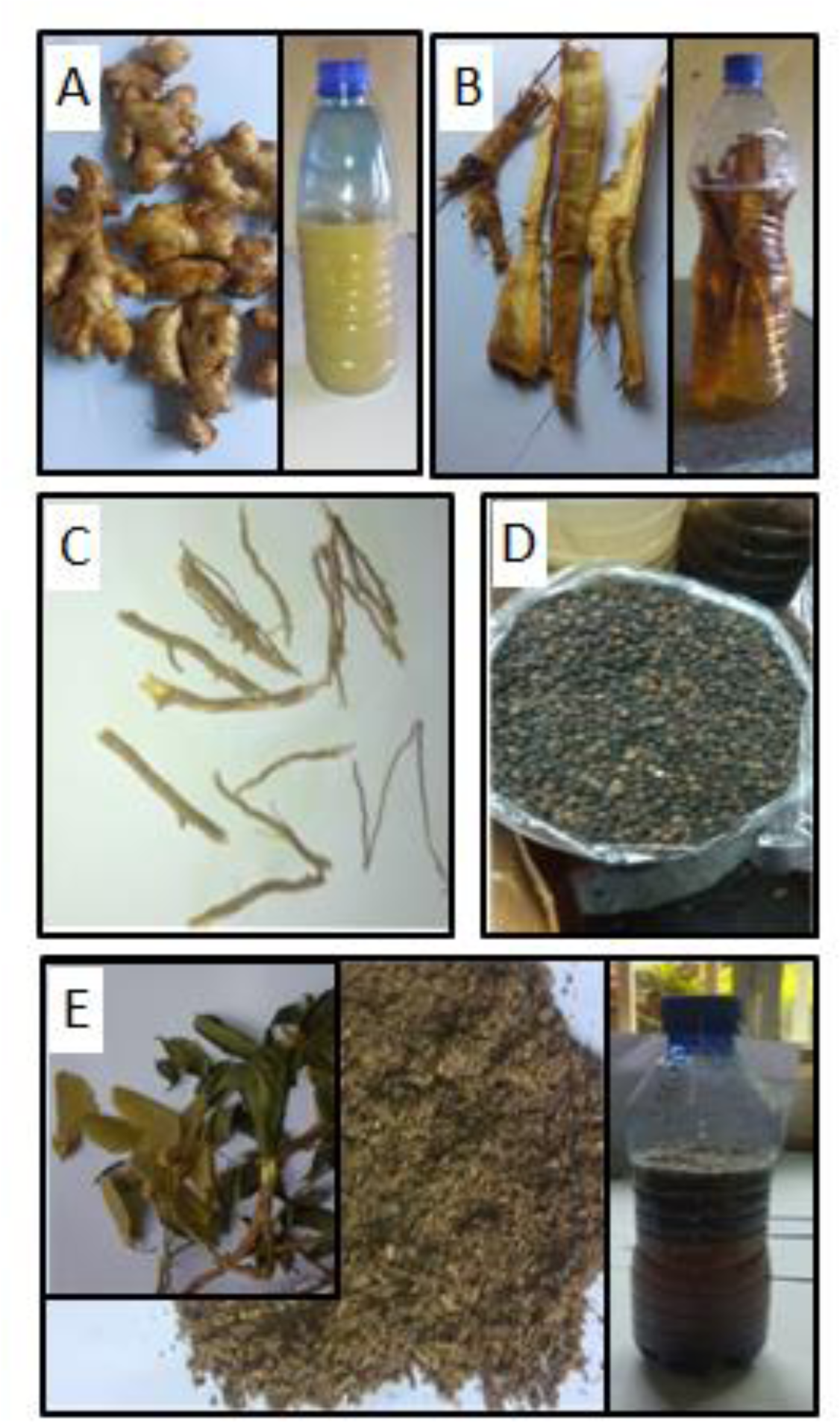
Traditional aphrodisiac products consumed by participants A. Tangawisi or ginger (Zingiber officinale) tubers (taken from the market), bottle with tangawisi decoction: B. Mujilanga (Mitragyna stipulosa): dried barks, bottle with infusion obtained from local alcohol mixed with dried Mujilanga barks C. Kafupa (Ocimum Sp) roots D. Nketu seeds (Piper nigrum) E. Mweyeye leaves, flowers and stem of Securidaca longepedunculata, (taken from a tree), bottle with infusion obtained from local alcohol with pounded bark of S. longepedunculata. (All photographs taken by PMO)

Five plants were reportedly consumed as aphrodisiacs (Table 3, Figure 1): **A**: Tangawisi *(Zingiber officinale),* **B**: Mujilanga *(Mitragyna stipulosa),* **C**: Kafupa (*Ocimum sp),* **D**: Nketu *(Piper nigrum*), **E:** Mweyeye *(Securidaca longepedunculata).* Among our participants, these agents were either consumed orally or applied anally (Table 3). Seven participants among aphrodisiacs consumers declared combining PDE5i with aphrodisiac products.

**Table 3:**
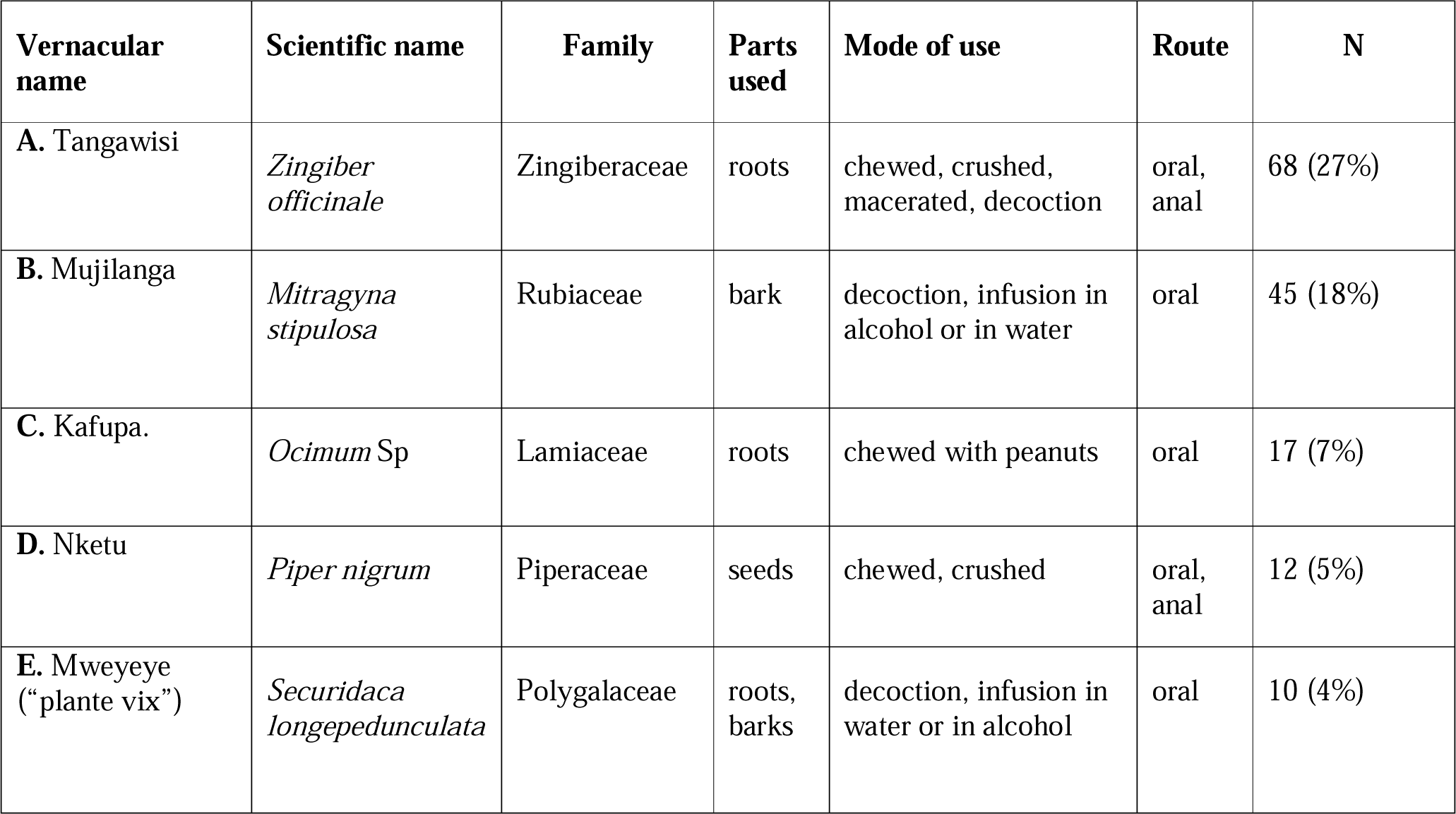
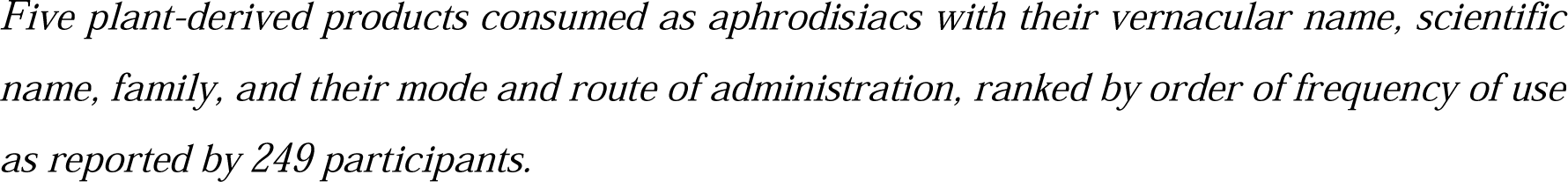
Description of reported plant-derived aphrodisiacs.

| Vernacular name | Scientific name | Family | Parts used | Mode of use | Route | N |
| --- | --- | --- | --- | --- | --- | --- |
| A. Tangawisi | *Zingiber officinale* | Zingiberaceae | roots | chewed, crushed, macerated, decoction | oral, anal | 68 (27%) |
| B. Mujilanga | *Mitragyna stipulosa* | Rubiaceae | bark | decoction, infusion in alcohol or in water | oral | 45 (18%) |
| C. Kafupa. | *Ocimum* Sp | Lamiaceae | roots | chewed with peanuts | oral | 17 (7%) |
| D. Nketu | *Piper nigrum* | Piperaceae | seeds | chewed, crushed | oral, anal | 12 (5%) |
| E. Mweyeye (“plante vix”) | *Securidaca longepedunculata* | Polygalaceae | roots, barks | decoction, infusion in water or in alcohol | oral | 10 (4%) |

*Five plant-derived products consumed as aphrodisiacs with their vernacular name, scientific* *name, family, and their mode and route of administration, ranked by order of frequency of use* *as reported by 249 participants.*

Two consumed aphrodisiacs (*Securidaca longepedunculata* and *Mitragyna stipulosa*) were taken with alcohol. The reported consumption of aphrodisiacs was 1.6-fold more prevalent among taxi-drivers compared to miners (chi square test= 20.8, p<0.001), and the types of aphrodisiacs used also differed between the two groups (Figure 3).

**Figure 3:**
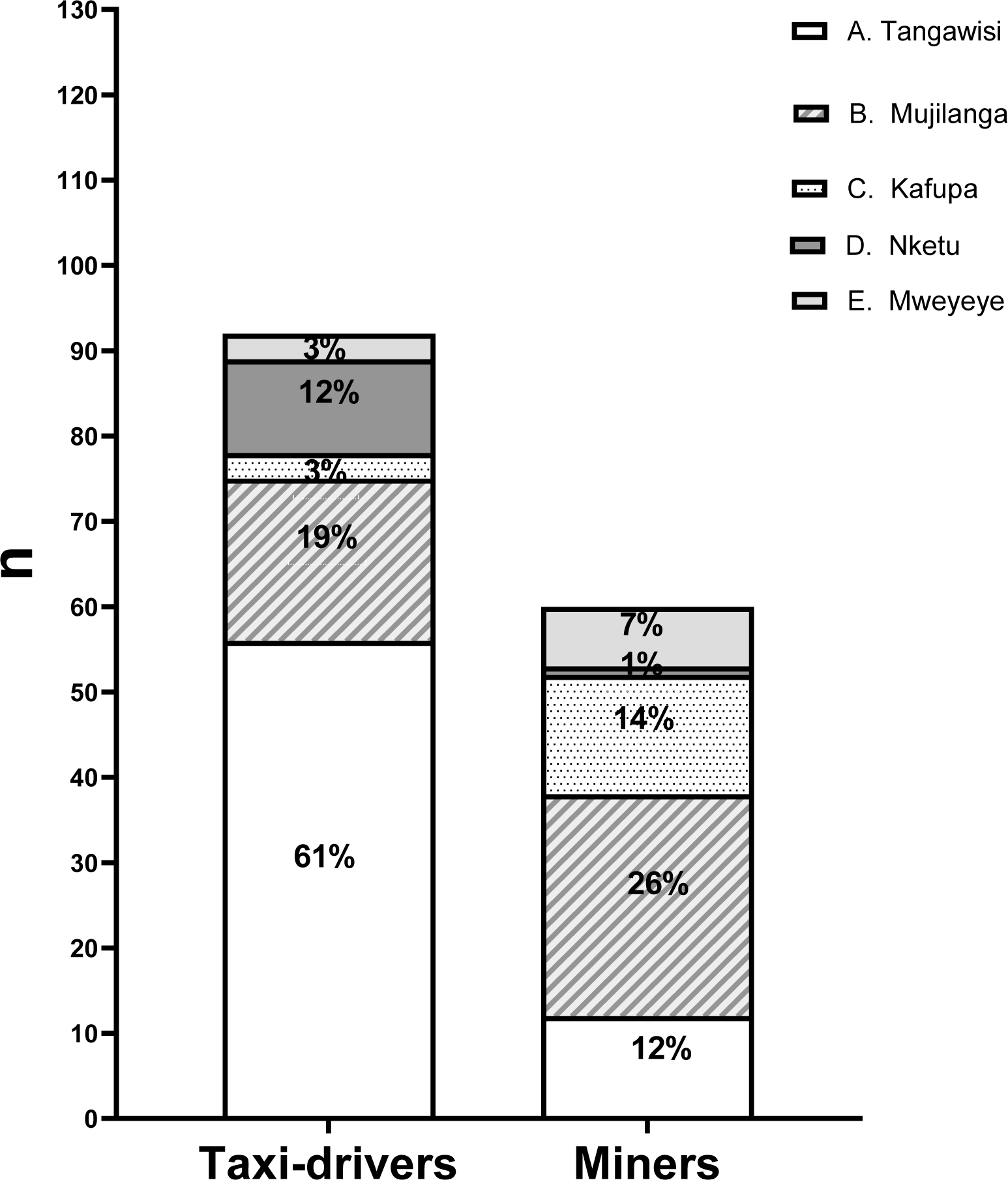
Prevalence of aphrodisiacs consumption among 127 collective taxi-drivers and 122 miners Proportion of men reporting consumption of five specific plant-based aphrodisiacs at least once a week as assessed in a survey of 127 collective taxi-drivers and 122 miners. Significantly more taxi-drivers than miners reported using aphrodisiacs, and the type of products also differed between the groups (chi square test= 20.8, p<0.001)

Of note, aphrodisiacs were also consumed by 54% of men without complaints of ED, whilst 36% of men with complaints of ED did not report taking aphrodisiacs.

### 3.3. Laboratory results

No differences were found between aphrodisiac consumers and non-consumers regarding serum levels of total testosterone, free testosterone, SHBG, or LH (Table 4).

**Table 4:** Hormones.

| Hormones | Regular consumers n = 137 | Non-consumers n = 72 | P-value |
| --- | --- | --- | --- |
| Total Testosterone (ng/dL) | 461 (367 – 593) | 472 (320 – 577) | 0.87 |
| Free Testosterone (ng/dL) | 10 (8 – 13) | 10 (8 – 13) | 0.57 |
| SHBG (nmol/L) | 31 (19 – 43) | 32 (20 – 42) | 0.50 |
| LH (IU/l) | 6 (5 – 8) | 6 (5 – 8) | 0.95 |

*Values reported are medians with interquartile range (IQR). P-value by Mann Whitney U-test*
*SHBG: Sexual Hormone Binding Globulin*
*LH: Luteinizing Hormone*

Table 5 presents the biochemical parameters in blood of 137 aphrodisiac consumers and 72 non-consumers. Overall, regular consumers exhibited similar or slightly *better* values for serum markers of hepatic or renal injury than non-consumers. However, total cholesterol, triglycerides, HDL and LDL were significantly higher among consumers than non-consumers. After adjusting for age, tobacco smoking, alcohol consumption, high blood pressure, overweight and obesity, aphrodisiacs consumption was significantly associated with LDL [adjusted odds ratio (aOR) 2.1 (95% confidence interval 1.1 – 3.8), P=0.01], and with total cholesterol [aOR 3.3 (1.6 – 6.8), P=0.001].

**Table 5:**
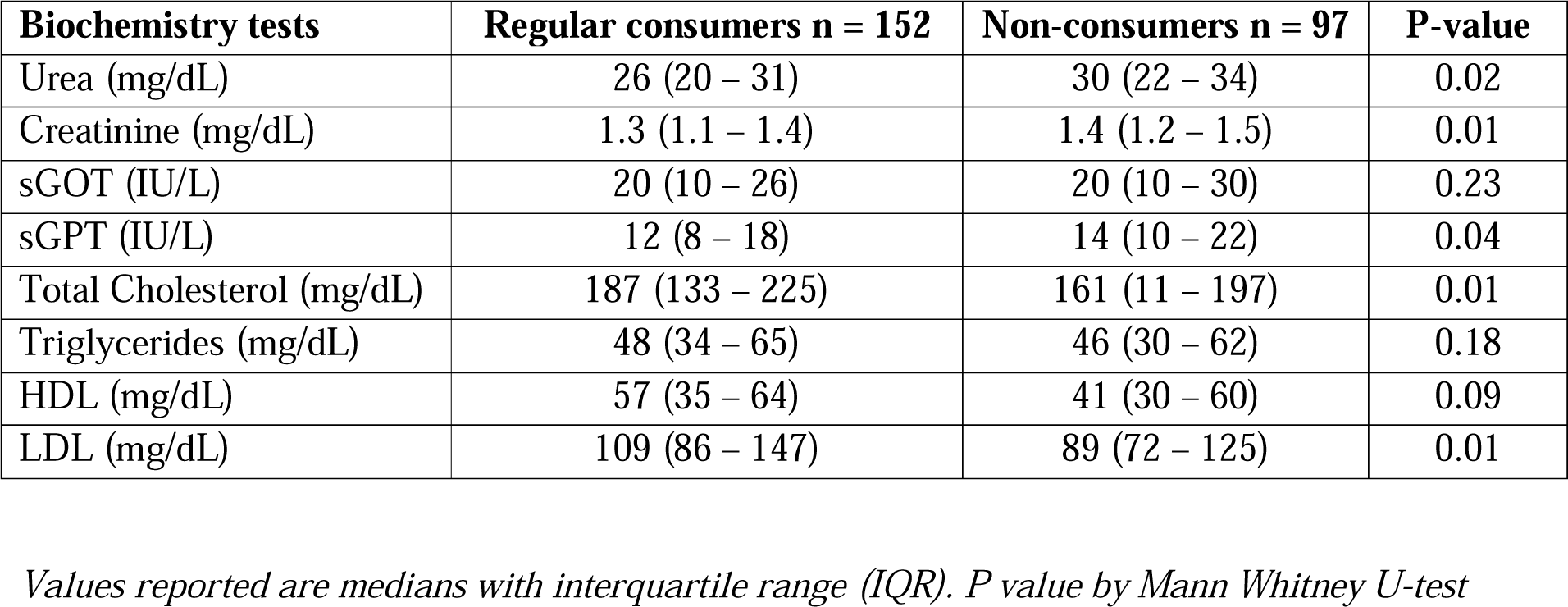
Biochemistry results.

No specific aphrodisiac product stood out in a non-parametric analysis of variance (Kruskall-Wallis) with comparisons made between non-consumers and consumers of each of the five types of aphrodisiacs (Supplementary table 1).

### 3.4. Qualitative results

All participants in the qualitative study were (by design) regular users of natural aphrodisiacs. The eight collective taxi-drivers included in the qualitative study were somewhat older (36±6 y) than the eight artisanal miners included (34±3 y), but artisanal miners had been in their jobs longer (12±4 y) than the collective taxi-drivers (9±4 y). Both groups contained men with and without ED, as defined by the IIEF questionnaire; 5 taxi-drivers had mild ED (score 19 – 24) compared to 3 miners including 2 with mild ED (score 19 – 24) and 1 with moderate ED (7 – 12).

In both groups, erectile dysfunction was described as a sign of loss of male identity. Participants considered erection as the main man’s attribute.

*“I often take these aphrodisiac plants to always maintain “Nguvu”* [sexual performance], *to always remain the head of my house and always give orders to my wife. Otherwise if you become sexually weak, it is very bad for the home and you become unhappy”* (T3).

“*The value of a man is a good erection* (…) *we consume these plants because our job weakens us; without the consumption of these stimulants we will not be real men in our respective houses (…) you know*, *once you are able to satisfy sexually your wife, all other things do not have a value, even if you do not have a job*” (M1).

“*Doctor, I can tell you, if you start experiencing difficulties with erection your mother in law will be the first person to be told about it. (…) let me tell you the truth, when we meet during breaks we often talk about our sexual life, we have common issues (…). Our partners are complaining about a decrease in our sexual power”* (M2).

Miners were afraid of losing their relationship with their wives. Most of them believed that being a man implies the ability to maintain one’s erection for penetrative sex, and to satisfy a woman, and that this is very crucial for a better consideration in the society.

“*I sometimes feel guilty when I can’t satisfy my wife sexually (…) but, as you know, the hip is the pillar of the house. Otherwise if your hip no longer works well, you become a dead man and your home will be doomed or your wife will be at the mercy of whoever can satisfy her*” (M1).

Among miners, interviews revealed a wide belief that the arduous nature of their work combined with some toxic agents present in the mined minerals, affected their sexual health:

*“Doctor, if you are doing a hard work your anus will be permanently open and this condition can’t allow a man to have a better sexual life”* (M2).

Some were desperate as they lost confidence, they also believed that their relationships were already affected, and they thought, they were cheated by their partners.

“*Almost all our wives complain about our sexual performance. It is believable that those who are smart already have friends with whom they have fun”* (M3).

Repeated failed sexual activities due to erectile dysfunction were the main reason that led most miners to seek an ultimate solution to recover respect as a man. The best solution was to look for products that could allow them to satisfy their partners. These power products were first taken to experiment with having sex with an extra-marital partner to ensure that they would work.

Among taxi-drivers, the prolonged sitting position during their job and the chronic low back pain caused by shocks due to poor road conditions were believed to be risk factors that could lead to ED, which could be prevented by taking preventive measures.

“*Doctor, do you know that as we spend hours being seated, this can compress our anus along with “Mishipa”* [vessels and nerves] *that supply the penis (…). These factors are capable to affect our future sexual health (…) that is why we must maintain our backs and anus by taking power products after work to prevent any unwanted sexual issue”* (T1).

“*But when we take them regularly, we feel like the hip becomes more flexible. This is why we trust all these herbal teas that these women sellers bring us because they help us prevent the worst in our couples*” (T2).

## 4. Discussion

This mixed methods study was prompted by our observations of a high prevalence of erectile dysfunction among mineworkers and its association with a high consumption of local aphrodisiacs [10]. In two large groups of male workers, i.e., artisanal miners and collective taxi-drivers, we assessed sexual function by means of a validated questionnaire, inquired about the types and patterns of use of aphrodisiacs, and explored their possible adverse effects on liver and kidney functions, and we then interviewed a subsample of participants to explore the main reasons for using aphrodisiac plants. To our knowledge, this is the first study of its kind in sub-Saharan Africa.

### General considerations

As in our previous study [10], we found a high prevalence of questionnaire-derived ED (IIEF-EF score<25) among (relatively young) miners: 34% in the present study, among diggers from three artisanal mines in Kolwezi, and even 47% in our previous study, among mineworkers of similar age but recruited from a larger geographical area and a broader range of mining jobs. However, the prevalence of ED differed considerably between the controls of the present study (collective taxi-drivers) and those of the previous study (bakers): 55% *versus* 17%, respectively. The very high prevalence of reported ED and use of aphrodisiacs among the taxi-drivers, compared to miners, was unexpected and cannot be attributed to older age or other characteristics. We have no firm explanation for the differences in ED prevalence between our two control populations. In our previous survey [10], we had concluded from a mediation analysis that erectile dysfunction among miners was partly explained by poor quality of marital relationships, possibly due to long time spent working away from home. This was possibly also the case for our taxi-drivers, but we were not able to assess this aspect in the current study, in which we did not assess the quality of the partner relationship. It is also conceivable that responses, even to validated questionnaires, are heavily influenced by perceptions of sexual health and masculinity, which may substantially vary between groups of workers, even within categories with similar socio-economic status.

However, the aim of the present study was not so much to investigate the prevalence of ED in different occupational groups, but to focus on the use of aphrodisiacs. Interestingly, although ED (mild-to-moderate and moderate) was 2-fold more prevalent among consumers of aphrodisiacs than among non-consumers (Table 2), aphrodisiacs were also consumed by men without complaints of ED, and not all men with complaints of ED reported taking aphrodisiacs.

As in many African cultures, aphrodisiac products are mostly made from plants and they are widely consumed in main cities of DR Congo. The overall prevalence of self-reported regular consumption of aphrodisiacs found in our study (61%) was somewhat higher than the 53% reported among men of the same age in Ghana [18]. This is consistent with a study in Kinshasa in which 59% of men aged from 35 to 49 years reported using aphrodisiacs [19]. Also, the prevalence found in our study was 4-fold higher than that reported in another study from Ghana [20]. In our study population, aphrodisiacs could be purchased easily at their working places, including bus stops and mines. Moreover, advertisements for “power products” including energy drinks [21], possibly contribute to the high prevalence of aphrodisiacs consumption in our region. Aphrodisiacs were often consumed mixed with alcohol or together with alcoholic beverages, therewith corroborating a study from Ghana [18]. Our aphrodisiacs consumers often said that drinking alcoholic beverages improved the effect of aphrodisiac products. In small doses alcohol can improve erections and increase desire due to its anxiolytic effects [22]. However, chronic alcoholism may cause hypogonadism and neuropathy, both of which may interfere negatively with erectile function [23].

### Properties of the aphrodisiacs consumed in southern Katanga

Available studies on aphrodisiacs have identified different mechanisms through which they could act [1]. These mechanisms include NO and cGMP signalling, NOS expression, increase (LH, testosterone and FSH) or decrease (prolactin) in hormone levels, co-ordination of central, sympathetic and parasympathetic nervous system [3].

In general, scientifically valid information on these plants’ pharmacological properties and effects on sexual performance are limited, and virtually non-existent for humans. Results from a systematic review of medicinal plants used for sexual dysfunction in sub-Saharan Africa demonstrated that out of the 209 species that are used traditionally, only 48 were validated scientifically, while no information was found for the remaining 161 [24].

Some experimental studies undertaken in the field of herbal medicine have identified bioactive compounds responsible for erection or for libido in some of these plants, but the human relevance of these findings remains unknown, as well as their harm thresholds.

Here, we summarize the (limited) existing literature on the aphrodisiac plants reportedly used by the participants in this study.

#### A. Tangawisi (Zingiber officinale)

Ginger belongs to the Zingiberaceae family and is native to south-eastern Asia. It is widely used as a spice and or medicine in many countries in the world. This appeared to be the most frequently used aphrodisiac among our participants, who reportedly took it either orally or anally in different forms. The tuber could be chewed, crushed, macerated, or it could be taken as a decoction. The tuber is widely used to treat ED in DR Congo [8].

With regard to its sexual stimulant properties, only animal studies are available. In an experimental study in male rats, Riaz [25] reported an increase in sperm quality and a significant decrease in plasma testosterone and LH levels. However, in a study using oral gavage of ginger, Afzali and Ghalehkandi [26] reported an increase in semen testosterone levels in male rats in comparison to control rats. Banihani [27] suggested that ginger increases testosterone by enhancing the activity of antioxidant enzymes, normalizing blood glucose, increasing blood flow in the testes, increasing testicular weight, and recycling testosterone receptors.

#### B. Mujilanga (Mitragyna stipulosa)

Mujilanga belonging to the family Rubiaceae, is indigenous to Africa and is used in traditional medicine to heal various diseases. It is widely used by both men and women to treat gastrointestinal diseases. In men, it is used for sexual purposes, with either bark or roots of the plant being mixed with alcohol or water and then taken as an infusion or a decoction.

Experimental studies have reported trypanocidal [28], anti-inflammatory and cardiovascular activities [29], but clear and convincing literature about mechanisms related to sexual dysfunction treatment in animals or humans is not available.

#### C. Kafupa (*Ocimum* sp.)

Kafupa is a name given to various plants; here it seems that it was *Ocimum vanderystii*, a species of the family of Lamiaceae (basil) that is native of Congo, DR Congo, Zambia and Angola. In the Copperbelt region, the plant is indicative of a rich copper deposit [30]. Consumers often chew roots with peanuts to enhance erectile function. No scientific information seems available about this plant’s pharmacologic properties.

#### D. Nketu (Piper nigrum)

The pepper plant is a flowering vine in the family of Piperaceae, cultivated for its fruits (the peppercorn), which is usually dried and used as a spice and seasoning. Black pepper is the world’s most traded spice and one of the most commonly used spices added to cuisines around the world. Ground, dried, and cooked peppercorns have been used since antiquity, both for flavour and as a traditional medicine, and these different uses were also reported by consumers in the present study. They described using pepper as an aphrodisiac through the anus or orally by chewing it.

An experimental study has suggested different mechanisms of action through which black pepper may enhance sexual function in men. First, piperine, one of its components, may enhance sexual function in men given its inhibitory effect on testosterone 5”-reductase resulting in higher testosterone levels [31]. Secondly, fatty acids in black pepper, i.e., auric acid, myristic acid and palmitic acid, affect the secretion and metabolism of androgens. Lastly, *Piper nigrum* is known to contain zinc that increases serum levels of sex hormones including testosterone.

#### E. Mweyeye (Securidaca longepedunculata)

Mweyeye is an herb belonging to the family of Polygalaceae (“violet tree)”. Participants in the present study reported consuming this plant’s bark after decoction or infusion in alcohol or simply water for about 48 hours. According to local sellers, *“the powder from pounded barks of the plant can be applied on the root and the body of the penis 45 minutes to one hour before intercourse to improve one’s erection. Mweyeye’s powder is widely sold in Zambia, South Africa, and in almost the entire eastern part of Africa under the name* ‘*Congo dust’ or* [in Swahili] ‘*Vumbi la Congo’ (…)”*.

Chloroform extracts of the root bark of this plant have been shown in vitro to induce relaxation of corpus cavernosum smooth muscle, almost as effectively as sildenafil [32].

In conclusion, although these natural plant-derived aphrodisiacs are widely consumed, little to no reliable information seems to exist about their pharmacological or toxicological properties for humans subjects. None of the plants reportedly used as aphrodisiacs in the present study are mentioned in a review of “scientifically proven herbal aphrodisiacs” ^1^.

### Comparisons between men consuming aphrodisiacs and men not consuming aphrodisiacs

We did not find differences between the two groups regarding serum levels of sex hormones. We also did not pick up complaints of adverse effects, such as priapism, during our survey. Total cholesterol and low-density lipoprotein were significantly higher among aphrodisiacs consumers than among non-consumers. We do not have explanations for these results but the higher prevalence of tobacco smoking found among our aphrodisiacs consumers could explain high lipid levels, since tobacco smoking is known to play a role in the occurrence of dyslipidemia [33,34]. Hypertension also appeared to be more prevalent among regular users of aphrodisiacs, possibly also in relation to smoking.

It is known that plants containing pyrrolizidine alkaloids may cause hepatic veno-occlusive disease [35] and acute renal failure is a common complication in poisoning by plants or mushrooms [35]. However, serum markers did not reveal hepatotoxicity or nephrotoxicity among consumers compared to non-consumers. Admittedly, our cross-sectional study was insensitive to document hepatoxicity or nephrotoxicity among these relatively young and active men. We also did not detect significant differences among the different aphrodisiac agents used, although the numbers of subjects were low for some compounds. Another limitation is that we did not attempt to evaluate the quantities of aphrodisiacs taken by the participants. Studies should be conducted with designs allowing comparisons of effects within individuals, i.e., before and after consuming well defined compounds of interest.

### Reasons for consuming aphrodisiacs as emerging from our qualitative study

Based on the rapid saturation achieved in the information obtained from the interviews, we may conclude that perceptions about masculinity were largely similar for miners and taxi-drivers.

However, the themes that emerged as reasons for taking aphrodisiacs differed between the two groups: taxi-drivers resorted to aphrodisiacs mainly for preventive purposes, as illustrated by the following quote from a taxi-driver:

“*When we take them regularly we feel like the hip becomes more flexible. This is why we trust all these herbal teas that these women sellers bring us because they help us prevent the worst in our couples*” (T2)

In contrast, miners took aphrodisiacs as a treatment to support failing erections, as evoked by one miner:

“*I haven’t been able to make love for almost 3 years without using tonics and especially because I’m afraid that my wife will leave me because we’re all still young*” (M6)

In Congolese cultures, there is a wide belief that a man must be dominant and possessive: men need to satisfy sexually women through penetrative sex regardless of his age, his mental state or his medical condition. Being a man implies maintaining one’s erection long enough for penovaginal sex, and failure could result in masculinity loss, and/or infidelity of the wife. Men’s ejaculation is an indicator of orgasm. Early ejaculation is an indicator of sexual dysfunction. Different Swahili expressions commonly used like: *Ule ni mwenye kufa tshini* (that one is dead down) to mean someone who is weak sexually; *Ule ni djogo* (that one is a rooster) to mean someone who ejaculates early; *Ule eko na ka bamiya* (that one has okra) to mean someone who has a small penis (like the legume).

The aforementioned cultural beliefs and considerations in Congo, as in most African countries, could play an important role in the widespread use of aphrodisiacs, and also contribute to a continuous search of an “ultimate” aphrodisiac for a perfect sexual life and a good reputation as a “real man” in society.

Our qualitative study was limited by the relatively low number of participants (even though we achieved saturation of the information), but we recognize that we could have obtained additional relevant information on the men’s motivations to consume aphrodisiacs if we had included non-users (with and without complaints of ED), as well as traditional healers advising or producing aphrodisiacs, or sellers of these products. More importantly, we acknowledge a major limitation in that our study did not involve women, wives and other partners of miners and taxi-drivers. This will be done in future studies.

## Conclusions

In a male worker population with high prevalence rates of reported ED, we found a high prevalence of regular consumption of various plant products used as natural aphrodisiacs. The widespread use of these herbal remedies is based on cultural beliefs about masculinity and male sexuality but the reasons for resorting to them appeared to differ between miners, who needed products to support failing erection, and taxi-drivers, who wanted them to prevent sexual problems. We found no evidence of nephrotoxicity, hepatotoxicity or hormonal disturbances in men using these compounds. Nevertheless, more ethnobotanical studies of plants consumed as aphrodisiacs, focussing on isolation and rigorous identification of the active constituents, are needed to provide the poor population in DRC, and globally, with correct information to protect them against possible toxic effects of traditional ‘remedies’ intended to improve or protect sexual function.

## Supporting information

Comparison of biochemical parameters according to specific types of consumed aphrodisiacs

## Data Availability

The data for this study will be made available upon reasonable request to the corresponding author.

## Acknowledgments

We thank the participants for their participation, and their representatives, as well as local authorities, for facilitating or allowing this study.

## Competing interest

The authors declare they don’t have conflicts of interest.

The data for this study will be made available upon reasonable request from the corresponding author.

## Author contributions

PMO, PE and BN conceived and designed the study. CLBN supervised the field study. PMO, GKM, JPK, TCK and TMM collected data in mining and among taxi-drivers. PMO cleaned and analysed the data under supervision of PE and BN. PMO wrote successive drafts of the paper, with input from JPK, TCK, GKM and TKK under the supervision of PE and BN. All authors approved the final version of the manuscript.

