## Supplementary material for "Natural aphrodisiacs consumption by male workers in the former Katanga province, DR Congo": Comparison of biochemical parameters according to specific types of consumed aphrodisiacs

**Supplementary table 1: Comparison of biochemical parameters according to specific types of consumed aphrodisiacs**

| **Biochemical parameters** | **Non-consumers**  **n= 72** | **A. Tangawisi**  **n= 61** | **B. Mujilanga**  **n =39** | **C. Kafupa**  **n =15** | **D. Nketu**  **n =12** | **E. Mweyeye**  **n = 8** | **P** |
| --- | --- | --- | --- | --- | --- | --- | --- |
| Urea (units) | 31 (22–34) | **24** (20–29) | **24** (20–30) | 25 (22–29) | 27 (22–31) | 25 (21–31) | **0.04^*^** |
| Creatinine | 1.4 (1.2 – 1.5) | **1.2 (1.1 – 1.2)** | 1.3 (1.2 – 1.4) | **1.31 (1.2 – 1.4)** | 1.4 (1.3 – 1.7) | 1.32 (1.2 – 1.4) | **0.01^*^** |
| sGOT | 21 (10 – 30) | 20 (10 – 26) | 14 (9 – 23) | 16 (10 – 23) | 15 (10 – 23) | 17 (8 – 25) | 0.39 |
| sGPT | 14 (10 – 22) | **11 (8 – 14)** | 13 (8 – 18) | 12 (9 – 16) | 12 (8 – 22) | 17 (8 – 25) | 0.28 |
| Total Cholesterol | 162 (111 – 196) | **190 (141 – 220)** | 184 (118 – 224) | 173 (129 – 194) | 184 (125 – 223) | 162 (116 –270) | 0.19 |
| Triglycerides | 47 (31 – 61) | 49 (44 – 75) | 48 (31 – 70) | 47 (34 – 58) | 47 (31 – 65) | 45 (31 – 72) | 0.65 |
| HDL | 41 (30 – ­60) | **58 (37 – 65)** | 58 (31 – 65) | 43 (33 – 60) | 43 (30 – 61) | 43 (29 – 62) | 0.29 |
| LDL | 90 (75 – 125) | **110 (89 – 143)** | 107 (86 – 149) | 109 (89 – 122) | 110 (91 – 132) | 110 (84 – 164) | 0.23 |

*Values reported are medians with interquartile range (IQR). P-value by Kruskall-Wallis test, significantly different values compared to non-consumers (Dunn’s post-hoc test) are indicated in bold	.*
